# BASIC: Bayesian Spiral Attention Classifier for Interpretable Medical Image Classification

**DOI:** 10.1101/2025.10.08.25337608

**Authors:** Abhinav Sagar

## Abstract

Accurate medical image classification is critical for early diagnosis and effective treatment planning. However, conventional deep learning models often fail to provide reliable uncertainty estimates, limiting their clinical applicability. In this study, we propose a novel Bayesian neural network architecture for medical image classification that integrates channel-wise and spatial attention mechanisms, including Squeeze-and-Excitation (SE) blocks and a novel Spiral Attention, to enhance feature representation. The proposed model employs a Bayes-by-Backprop approach in the fully connected layers to quantify both epistemic and aleatoric uncertainties, allowing for reliable prediction confidence estimation. We validate our approach on multiple benchmark datasets, including diabetic retinopathy, COVID-19 chest X-rays, skin lesion images, and gastrointestinal endoscopy images. Extensive experiments demonstrate that our method not only achieves high classification performance but also provides meaningful uncertainty estimates, improving interpretability and robustness in clinical decision-making. Additionally, qualitative analysis using Grad-CAM visualizations highlights the model’s ability to focus on clinically relevant regions, further supporting its potential for real-world deployment.

## 1. Introduction

Medical image classification plays a pivotal role in early diagnosis, prognosis, and treatment planning across a wide range of clinical applications, including diabetic retinopathy detection, COVID-19 chest X-ray screening, skin lesion classification, and gastrointestinal endoscopy analysis. Deep learning models, particularly convolutional neural networks (CNNs) and transformer-based architectures, have demonstrated remarkable performance in these tasks due to their ability to automatically learn hierarchical feature representations from complex image data.

Despite these advances, conventional deterministic deep learning models suffer from a critical limitation: they often produce overconfident predictions and fail to quantify uncertainty, which can hinder their deployment in safety-critical clinical environments. Uncertainty estimation is essential to identify cases where the model may be unreliable and to provide actionable confidence information to clinicians.

To address these challenges, we propose a novel Bayesian neural network architecture that incorporates both channel-wise and spatial attention mechanisms. The network integrates Squeeze-and-Excitation (SE) blocks and Spiral Attention modules within residual blocks to capture informative features across channels and spatial locations effectively. The fully connected layers are replaced with Bayes-by-Backprop layers, allowing the model to estimate both epistemic and aleatoric uncertainties. This enables robust predictions and confidence-aware decision-making in medical image classification tasks.

We validate the proposed method on multiple benchmark medical image datasets, including diabetic retinopathy, COVID-19 chest X-rays, skin lesion images, and gastrointestinal endoscopy images. Extensive experiments demonstrate that our approach achieves high classification performance while providing meaningful uncertainty estimates, improving interpretability, and enhancing clinical reliability. Moreover, Grad-CAM visualizations confirm that the attention mechanisms enable the model to focus on clinically relevant regions, providing additional interpretability and insight into its decision-making process.

To further investigate the interpretability of our framework, we visualize the GradCAM-based attention maps along with uncertainty quantification on the Diabetic Retinopathy as shown in Fig. 1. The highlighted regions in the attention maps correspond to discriminative areas that strongly influence the model’s decision-making process. In the case of diabetic retinopathy, the model primarily focuses on microaneurysms and hemorrhagic regions. The uncertainty values provide an additional layer of interpretability by indicating how confident the model is in its predictions for each sample. Together, these visualizations demonstrate that our model not only achieves high predictive performance but also offers trustworthy explanations, which are critical in medical imaging applications where interpretability and reliability are paramount.

**Figure 1.**
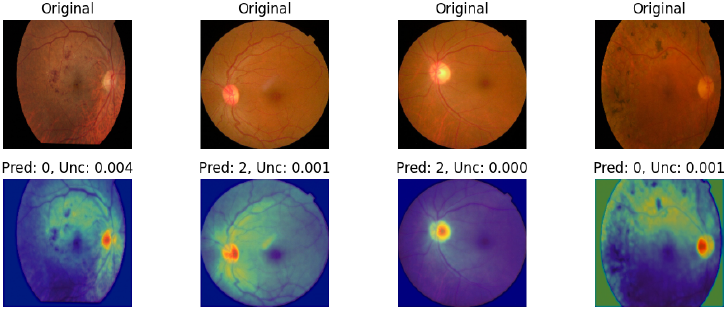
Illustration of GradCAM-based attention maps along with predicted labels and uncertainty quantification using the diabetic retinopathy dataset.

The three main contributions of our paper are as follows:

- We propose a Bayesian neural network for medical image classification that incorporates channel-wise (SE) and spatial (Spiral) attention mechanisms to improve feature representation and robustness.
- We integrate Bayes-by-Backprop layers to quantify both epistemic and aleatoric uncertainties, providing confidence-aware predictions suitable for clinical applications.
- We demonstrate the effectiveness of the proposed approach across multiple medical image datasets, showing superior performance, meaningful uncertainty estimation, and enhanced interpretability via Grad-CAM visualizations.

## 2. Related Work

Medical image classification has seen remarkable progress due to deep learning, with approaches spanning CNN-based, transformer-based, and hybrid architectures. In addition, uncertainty-aware methods have been increasingly applied to enhance reliability in clinical applications.

### 2.1. CNN-Based Medical Image Classification

CNNs have been foundational in medical imaging, offering strong hierarchical feature extraction. Classic architectures such as ResNet He et al. (2016), DenseNet Huang et al. (2017), EfficientNet Tan and Le (2019), and RepVGG Ding et al. (2021) have been widely applied for tasks including retinal fundus classification Fang et al. (2019); Asia Pacific Tele-Ophthalmology Society (APTOS) (2019), gastrointestinal disease detection Pogorelov et al. (2017), skin lesion analysis Codella et al. (2019), and chest X-ray classification Shastri et al. (2022); Okolo et al. (2022).

Recent work has explored attention-enhanced CNNs, including ResGANet Cheng et al. (2022), BARF Abdar et al. (2021), and Hercules Abdar et al. (2022), to focus on salient regions and improve feature fusion. Techniques such as SE blocks Hu et al. (2018) and spatial attention further refine channel-wise and spatial feature representations, enabling more accurate diagnosis.

### 2.2. Transformer-Based Medical Image Classification

Vision Transformers (ViTs) Dosovitskiy et al. (2020) and their variants have shown strong performance by modeling long-range dependencies. MedViT Manzari et al. (2023), TransMed Dai et al. (2021), MedFormer Chowdary and Yin (2024), and Multi-scale Transformer models Hu et al. (2025) demonstrate the effectiveness of pure transformer and hybrid transformer-CNN architectures in multi-modal and generalized medical image classification. These models, while powerful, often require large datasets or pre-training on natural images Xie and Richmond (2018); Xue et al. (2019); Huix et al. (2024).

### 2.3. Hybrid CNN-Transformer Architectures

To balance CNN efficiency and transformer global context modeling, hybrid architectures have emerged. Examples include CTransCNN Wu et al. (2023), EFFResNet-ViT Hussain et al. (2025), DBCvT Li et al. (2024a), and MobileViT-based medical classification networks Mehta and Rastegari (2021). These models combine convolutional feature extraction with transformer attention to handle both local and global patterns effectively.

### 2.4. Uncertainty-Aware and Robust Classification

Model uncertainty is crucial in medical applications. Bayesian and uncertainty-aware methods, such as Monte Carlo dropout Gal and Ghahramani (2016), BARF Abdar et al. (2021), Hercules Abdar et al. (2022), BayTTA Sherkatghanad et al. (2025), and dual-uncertainty estimation Ju et al. (2022), provide predictive confidence for safer clinical deployment. Techniques like balanced-mixup Galdran et al. (2021), co-training with noisy labels Xue et al. (2022), and model calibration Rajaraman et al. (2022); Liang et al. (2020) enhance robustness against imbalanced or noisy datasets.

### 2.5. Data Augmentation and Pre-training

Effective data augmentation Hussain et al. (2018), pre-training strategies Xie and Richmond (2018); Zhou et al. (2023), and transfer learning Raghu et al. (2019); Swati et al. (2019) have been widely used to mitigate limited annotated data in medical imaging. Advanced augmentation techniques, including multi-scale pyramid fusion Wen et al. (2025) and dynamic adaptive fusion Cai et al. (2025), further improve model generalization across diverse clinical datasets.

## 3. Methodology

### 3.1. Problem Definition

Medical image classification aims to automatically categorize medical images into predefined classes, such as disease types, severity levels, or anatomical structures. Formally, let 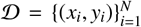 denote a dataset consisting of *N* medical images *x*_*i*_ ∈ ℝ^*H*×*W*×*C*^ and corresponding labels *y*_*i*_ ∈ {1, …, *K* }, where *K* is the number of classes. The goal is to learn a mapping function *f*_*θ*_ : ℝ^*H*×*W*×*C*^ → {1, …, *K*} parameterized by *θ*, which can accurately predict labels for unseen images *x*_test_. . Formally, the problem can be expressed as minimizing the expected classification loss ℒ over the data distribution *P*(*x, y*):

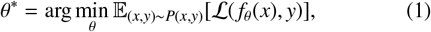

where ℒ may incorporate cross-entropy loss, uncertainty penalties, or class-balancing terms depending on the model design. The ultimate objective is a robust, accurate, and generalizable classifier capable of handling diverse medical imaging datasets.

### 3.2. Network Architecture

The proposed image classification network is designed to handle multi-class medical image datasets with uncertainty estimation. It integrates a convolutional residual backbone with attention mechanisms and a Bayesian classification head.

#### Stem

The network begins with a convolutional stem that consists of a 3 × 3 convolution followed by batch normalization and ReLU activation. This stem converts the input image into an initial set of feature maps.

#### Residual Blocks with Attention

Four sequential residual blocks process the feature maps. Each block contains two 3 × 3 convolutional layers with batch normalization and ReLU activation, combined with a skip connection. Two types of attention mechanisms are integrated into each block:

- **Squeeze-and-Excitation (SE) Block:** Applies global average pooling and a small fully connected network to compute channel-wise attention, scaling each channel adaptively.
- **Spiral Attention:** Aggregates features along multiple spiral paths radiating from the feature map center, enhancing spatial feature representation with a residual connection.

Downsampling is applied selectively in the residual blocks using a stride of 2, enabling hierarchical feature extraction.

#### Global Pooling and Dropout

After the residual blocks, the feature maps are pooled to a single vector using adaptive average pooling. Dropout is applied to reduce overfitting.

#### Bayesian Classification Head

The pooled features are passed through a Bayesian linear layer, which models weight uncertainty using a Gaussian variational posterior. The network can optionally produce aleatoric uncertainty by outputting both mean and variance for each class.

#### Forward Pass

For an input image *x*, the forward pass can be summarized as:

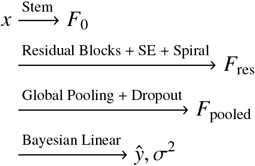

where *ŷ* represents the predicted class probabilities and *σ*^2^ encodes predictive uncertainty. The network supports Monte Carlo Dropout inference to estimate epistemic uncertainty.

The two novel contributions of our work, namely Spiral Attention and Bayesian Linear Layer, are described in the following below:

#### Spiral Attention

Spiral Attention is designed to enhance spatial feature representation by aggregating information along spiral trajectories across the feature map. For an input feature map of size *B* × *C* × *H* × *W*, a set of offsets is generated along multiple spiral spokes radiating from the center of the feature map. Each offset defines a sampling location, and bilinear interpolation is used to extract feature values at these locations. The outputs of all spiral paths are averaged and passed through a 1 × 1 convolution to generate a residual feature map:

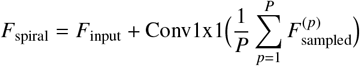

where *P* is the total number of spiral sampling points. This mechanism allows the network to capture long-range dependencies and directional feature patterns in a structured manner.

#### Bayesian Linear Layer

The Bayesian Linear Layer models the uncertainty in the classification head weights using a Gaussian variational posterior. Each weight and bias is parameterized by a mean *µ* and a variance *σ*^2^ (obtained via a softplus transformation of a learnable parameter *ρ*). During training, weights are sampled from the posterior distribution to compute the linear transformation:

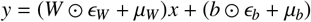

where *ϵ*_*W*_, *ϵ*_*b*_∼ 𝒩 (0, 1). The layer also computes the KL divergence between the posterior and a Gaussian prior to regularize the weight distribution:

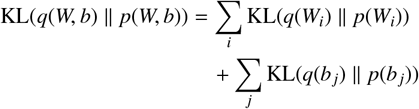

This Bayesian formulation allows the network to quantify epistemic uncertainty in predictions, which is critical for medical applications where reliable confidence estimates are required.

#### Overall Network Architecture

The proposed network architecture, illustrated in Figure 2, integrates advanced feature extraction and uncertainty modeling to improve classification performance. Panel (A) shows the overall network, starting with a convolutional stem that encodes the input image into feature maps, followed by a series of residual blocks augmented with Squeeze-and-Excitation (SE) and Spiral Attention modules to capture both channel-wise dependencies and spatial relationships along spiral trajectories. After feature extraction, a global average pooling and dropout layer prepare the representations for the classification stage. Panel (B) provides a closer look at the Spiral Attention module, which aggregates features from offset positions arranged in a spiral pattern, enabling the network to capture long-range spatial interactions efficiently. Panel (C) highlights the Bayesian Linear Layer, which applies Bayes-by-Backprop to model uncertainty in the final predictions, allowing the network to produce more reliable class probabilities while accounting for epistemic uncertainty in the model parameters. Together, these components form a robust, uncertainty-aware architecture suitable for challenging medical image classification tasks.

**Figure 2.**
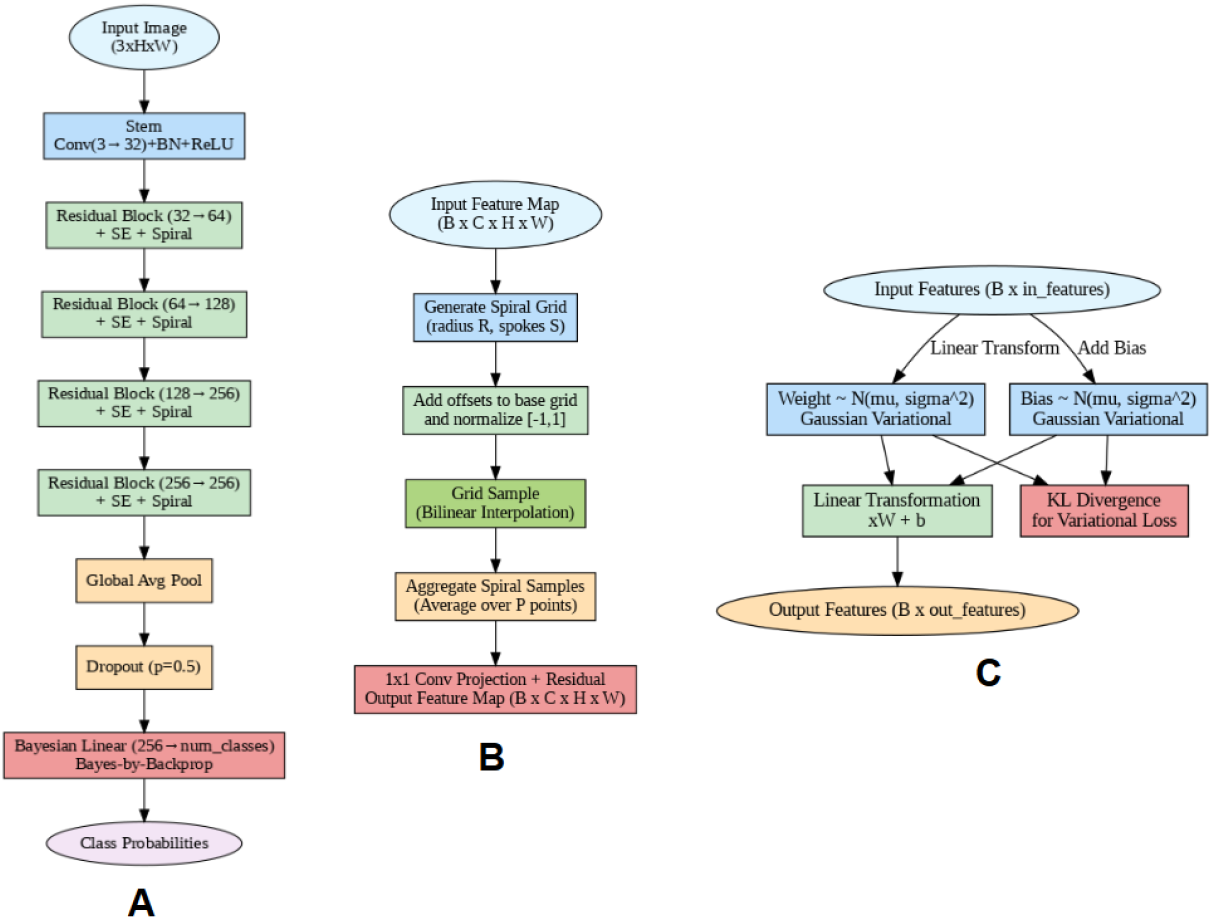
Illustration of the proposed network architecture. (A) Overall Network Architecture highlighting the stem, residual blocks with SE and Spiral Attention, global pooling, and Bayesian classifier. (B) Detailed view of the Spiral Attention module, showing how input features are aggregated along spiral offsets. (C) Detailed view of the Bayesian Linear Layer using Bayes-by-Backprop for uncertainty-aware classification.

### 3.3. Loss Function

The proposed model employs a composite loss function that combines cross-entropy, confidence calibration, class-adaptive weighting, feature margin regularization, and Bayesian regularization. This formulation encourages accurate predictions, calibrated confidence, and well-separated feature embeddings.

#### ACMLoss

The ACMLoss is defined as:

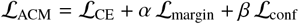

where:

- ℒ_CE_ is the standard cross-entropy loss between predicted logits and ground-truth labels.
- ℒ _conf_ = 1 − *p*_*t*_ is the confidence calibration term, which penalizes predictions with low probability *p*_*t*_ for the true class.
- ℒ_margin_ is the margin-based feature regularization applied to feature embeddings *f*_*i*_ and *f* _*j*_ of different classes:

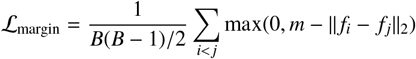

where *B* is the batch size and *m* is the margin hyperparameter.

- *α* and *β* are weighting factors for margin regularization and confidence calibration, respectively.
- Optionally, class-adaptive weighting adjusts ℒ_CE_ by inverse class frequency to address class imbalance.

#### Bayesian Regularization (KL Divergence)

For the Bayesian linear layer in the classifier, the KL divergence between the weight posterior *q*(*W, b*) and the Gaussian prior *p*(*W, b*) is computed:

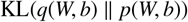

This term acts as a regularizer and encourages the weight distribution to remain close to the prior.

#### Total Loss

During training, the total loss combines the ACM-Loss and the Bayesian KL divergence, following the evidence lower bound (ELBO) formulation:

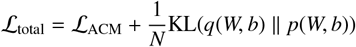

where *N* is the number of training samples. This ensures the network learns discriminative features while modeling predictive uncertainty.

## 4. Experiments

### 4.1. Dataset

The following datasets were used to test and compare the performance of our method:

1. **Covid-Pneumonia:** The COVID-Pneumonia dataset consists of chest X-ray images categorized into three classes: *Normal, COVID-19*, and *Viral*/*Bacterial Pneumonia*. The dataset includes 1,626 images of COVID-19 cases, 1,802 normal chest X-rays, and 1,800 pneumonia cases, providing a balanced distribution suitable for benchmarking medical image classification methods Shastri et al. (2022).
2. **Diabetic-Retinopathy:** The Diabetic Retinopathy (DR) dataset comprises high-resolution retinal fundus images categorized into five severity levels: *No DR, Mild, Moderate, Severe*, and *Proliferative DR*. The dataset contains 1,805 images with No DR, 370 Mild cases, 999 Moderate cases, 193 Severe cases, and 295 Proliferative DR cases, enabling comprehensive evaluation across the full spectrum of disease severity Asia Pacific Tele-Ophthalmology Society (APTOS) (2019).
3. **Kvasir:** The Kvasir dataset contains gastrointestinal (GI) tract endoscopic images annotated with both anatomical landmarks and pathological findings. It consists of 1,000 images for each of the following categories: dyed-lifted polyps, dyed-resection margins, esophagitis, normal cecum, normal pylorus, normal z-line, polyps, and ulcerative colitis, resulting in a balanced dataset of 8,000 images across eight classes Pogorelov et al. (2017).
4. **Skin Cancer:** The Skin Cancer dataset (HAM10000) consists of dermoscopic images labeled with nine types of skin lesions, covering both benign and malignant cases. Specifically, the dataset includes 130 images of actinic keratosis, 392 basal cell carcinoma, 111 dermatofibroma, 454 melanoma, 373 nevus, 478 pigmented benign keratosis, 80 seborrheic keratosis, 197 squamous cell carcinoma, and 142 vascular lesions. This diverse distribution across nine classes provides a challenging benchmark for skin cancer classification. Codella et al. (2019).

### 4.2. Implementation Details

#### Dataset and Preprocessing

All images were resized to 224 × 224 pixels and normalized with standard ImageNet statistics. Data augmentation included random horizontal flips during training. For cross-validation experiments, we employed 5-fold stratified splits to preserve class distributions, while for standard training, an 80/20 train-validation split was used.

#### Training

Models were trained using the Adam optimizer with an initial learning rate of 1 × 10^−4^. Batch sizes ranged from 4 to 8 depending on experiment setup. Training duration was set to 100 epochs for standard training and 100 epoch per fold in cross-validation experiments. Monte Carlo Dropout (30 samples) was used during inference to estimate epistemic uncertainty.

#### Evaluation Metrics

Performance was quantified using accuracy, precision, recall, F1-score, Matthews correlation coefficient (MCC), ROC-AUC, and Brier score. Calibration was assessed with the Expected Calibration Error (ECE) and reliability diagrams. Confusion matrices, ROC curves, and precision-recall curves were plotted for qualitative assessment. Grad-CAM was applied to visualize model attention maps.

#### Software and Hardware

The framework was implemented in Python 3.10 using PyTorch 2.1, torchvision, and scikit-learn. Training and evaluation were performed on a system equipped with an NVIDIA A100 GPU (CUDA 11.8), 32 GB RAM, and Intel Xeon CPU.

### 4.3. Comparison Approaches

To demonstrate the effectiveness of our proposed model, we compare it against several state-of-the-art approaches for medical image classification. The baselines are chosen to represent conventional CNN-based and transformer-based models.

#### 4.3.1. Quantitative Performance

We report the quantitative performance of our model and the baselines on four medical imaging datasets (Covid-Pneumonia Shastri et al. (2022), Diabetic Retinopathy Asia Pacific Tele-Ophthalmology Society (APTOS) (2019), Kvasir Pogorelov et al. (2017), and Skin Cancer Codella et al. (2019)). The following comparison approaches are considered: ConvMixer Trockman and Kolter (2022), DenseNet121 Huang et al. (2017), EfficientFormerL1 Li et al. (2022), EfficientNetB0 Tan and Le (2019), InceptionNextTiny Yu et al. (2024), MLPMixer Tolstikhin et al. (2021), MobileViTXXS Mehta and Rastegari (2021), MobileNetV3 Howard et al. (2019), RegionViT Chen et al. (2021), RepVGG Ding et al. (2021) Resnet50 He et al. (2016), ResNeXt101 Xie et al. (2017), ShuffleNetV2 Ma et al. (2018), SimpleViT Beyer et al. (2022), and VAN Guo et al. (2023).

The performance is evaluated using *accuracy, precision, recall, F1-score, AUC*, and *MCC* across all datasets. To assess uncertainty quantification, we further compute *predictive entropy, uncertainty, Brier Score*, and *expected calibration error (ECE)*.

Experimental results demonstrate that our proposed model consistently outperforms deterministic CNN and transformer baselines in terms of classification accuracy, while also providing well-calibrated uncertainty estimates.

Table 1 presents a comprehensive quantitative comparison across four diverse medical image classification datasets. Our proposed method consistently outperforms state-of-the-art CNN-, MLP-, and Transformer-based baselines in terms of accuracy, precision, recall, F1-score, AUC, and MCC. On the Covid-Pneumonia dataset, our approach achieves the highest overall performance, surpassing well-established backbones such as DenseNet121, EfficientNetB0, and ShuffleNetV2. Similarly, for the Diabetic Retinopathy dataset, our model provides substantial improvements in all major metrics, particularly in recall and F1-score, indicating robust sensitivity to disease-related patterns. On the Kvasir dataset, our method outperforms recent architectures by maintaining a superior balance between accuracy and MCC, highlighting its reliability in gastrointestinal image classification. Finally, on the more challenging Skin Cancer dataset, our model significantly surpasses competing methods, achieving a notable improvement of nearly 7–10% in accuracy and F1-score over strong baselines. Although our approach does not always achieve the lowest parameter count, FLOPs, or inference latency compared to ultra-lightweight models such as MobileNetV3 or ShuffleNetV2, the consistent improvements in predictive performance demonstrate the effectiveness of integrating residual-SE modules, Spiral Attention, and Bayesian uncertainty modeling into the classification framework.

**Table 1:** A: Quantitative comparison on the Covid-Pneumonia dataset. B: Quantitative comparison on the Diabetes dataset. C: Quantitative comparison on the Kvasir dataset. D: Quantitative comparison on the Skin Cancer dataset. The best values are highlighted in bold.

| A: Covid-Pneumonia Dataset |  |  |  |  |  |  |  |  |  |  |
| --- | --- | --- | --- | --- | --- | --- | --- | --- | --- | --- |
| Method | Accuracy ↑ | Precision ↑ | Recall ↑ | F1 Score↑ | AUC ↑ | MCC ↑ | Params ↓ | Flops ↓ | Time ↓ | FPS ↓ |
| ConvMixer Trockman and Kolter (2022) | 0.9744 | 0.9751 | 0.9747 | 0.9748 | 0.9959 | 0.9616 | 3.7084 | 3.8095 | 0.0109 | 91.5931 |
| DenseNet121 Huang et al. (2017) | 0.9742 | 0.9747 | 0.9745 | 0.9745 | 0.9971 | **0.9678** | 6.9507 | 2.8174 | 0.0176 | 56.6668 |
| EfficientFormerL1 Li et al. (2022) | 0.9583 | 0.9598 | 0.9588 | 0.9588 | 0.9954 | 0.9379 | 11.3869 | 1.3041 | 0.0069 | 145.7989 |
| EfficientNetB0 Tan and Le (2019) | 0.9738 | 0.9746 | 0.9742 | 0.9742 | 0.9967 | 0.9608 | 4.0108 | 0.4066 | 0.0105 | 95.2057 |
| InceptionNextTiny Yu et al. (2024) | 0.9692 | 0.9701 | 0.9694 | 0.9696 | 0.9961 | 0.9539 | 25.7479 | 4.1886 | 0.0092 | 108.8384 |
| MLPMixer Tolstikhin et al. (2021) | 0.9447 | 0.9457 | 0.9455 | 0.9455 | 0.9899 | 0.9171 | 5.1595 | 1.2656 | 0.0044 | 225.6782 |
| MobileViTXXS Mehta and Rastegari (2021) | 0.9671 | 0.9679 | 0.9678 | 0.9675 | 0.9966 | 0.9509 | 1.0121 | 0.2695 | 0.0117 | 85.1687 |
| MobileNetV3 Howard et al. (2019) | 0.9621 | 0.9637 | 0.9628 | 0.9625 | 0.9934 | 0.9439 | 1.5206 | **0.0578** | 0.0092 | 108.3404 |
| RegionViT Chen et al. (2021) | 0.9551 | 0.9558 | 0.9558 | 0.9555 | 0.9920 | 0.9327 | 12.1265 | 2.2312 | 0.0250 | 40.0033 |
| RepVGG Ding et al. (2021) | 0.9742 | 0.9745 | 0.9747 | 0.9745 | 0.9968 | 0.9614 | 10.2568 | 2.8521 | 0.0049 | 204.7226 |
| Resnet50 He et al. (2016) | 0.9635 | 0.9646 | 0.9641 | 0.9638 | 0.9957 | 0.9457 | 23.5079 | 4.0530 | 0.0107 | 93.8393 |
| ResNeXt101 Xie et al. (2017) | 0.9665 | 0.9666 | 0.9673 | 0.9667 | 0.9963 | 0.9500 | 42.1286 | 7.9742 | 0.0235 | 42.5254 |
| ShuffleNetV2 Ma et al. (2018) | 0.9749 | 0.9753 | 0.9752 | 0.9751 | 0.9965 | 0.9625 | 0.9194 | 0.1419 | 0.0072 | 138.3880 |
| SimpleViT Beyer et al. (2022) | 0.9306 | 0.9313 | 0.9320 | 0.9313 | 0.9856 | 0.8961 | **0.3469** | 0.0700 | **0.0027** | **376.6247** |
| VAN Guo et al. (2023) | 0.9742 | 0.9748 | 0.9745 | 0.9746 | 0.9969 | 0.9613 | 3.8432 | 0.8608 | 0.0101 | 99.0866 |
| Ours | **0.9749** | **0.9755** | **0.9755** | **0.9754** | **0.9978** | 0.9625 | 2.5618 | 3.3295 | 0.0632 | 15.8214 |
| B: Diabetic-Retinopathy Dataset |  |  |  |  |  |  |  |  |  |  |
| Method | Accuracy ↑ | Precision ↑ | Recall ↑ | F1 Score↑ | AUC ↑ | MCC ↑ | Params ↓ | Flops ↓ | Time ↓ | FPS ↓ |
| ConvMixer Trockman and Kolter (2022) | 0.7226 | 0.5197 | 0.5050 | 0.5040 | 0.8514 | 0.5783 | 3.7596 | 3.8609 | 0.0953 | 10.4885 |
| DenseNet121 Huang et al. (2017) | 0.7474 | 0.5729 | 0.5637 | 0.5611 | 0.8916 | 0.6195 | 6.9590 | 2.8960 | 0.0199 | 50.1820 |
| EfficientFormerL1 Li et al. (2022) | 0.7207 | 0.4685 | 0.4208 | 0.3845 | 0.8448 | 0.5741 | 11.3891 | 1.3095 | 0.0414 | 24.1817 |
| EfficientNetB0 Tan and Le (2019) | 0.6909 | 0.4698 | 0.4636 | 0.4631 | 0.8451 | 0.5314 | 4.0140 | 0.4139 | 0.0107 | 93.5750 |
| InceptionNextTiny Yu et al. (2024) | 0.6985 | 0.4988 | 0.4765 | 0.4799 | 0.8352 | 0.5419 | 25.7556 | 4.1982 | 0.0095 | 105.2280 |
| MLPMixer Tolstikhin et al. (2021) | 0.6966 | 0.4825 | 0.4793 | 0.4781 | 0.8382 | 0.5387 | 5.4226 | 1.3170 | **0.0043** | **230.3711** |
| MobileViTXXS Mehta and Rastegari (2021) | 0.7220 | 0.5124 | 0.4847 | 0.4779 | 0.8622 | 0.5756 | 1.0131 | 0.2731 | 0.0101 | 98.7030 |
| MobileNetV3 Howard et al. (2019) | 0.6980 | 0.4898 | 0.4830 | 0.4814 | 0.8363 | 0.5437 | 1.5230 | **0.0615** | 0.0091 | 109.6083 |
| RegionViT Chen et al. (2021) | 0.7130 | 0.5008 | 0.4666 | 0.4682 | 0.8447 | 0.5625 | 12.2360 | 2.2633 | 0.0600 | 16.6629 |
| RepVGG Ding et al. (2021) | 0.7032 | 0.5432 | 0.4969 | 0.5017 | 0.8401 | 0.5583 | 10.2591 | 2.8682 | 0.0418 | 23.9452 |
| Resnet50 He et al. (2016) | 0.7176 | 0.5363 | 0.5020 | 0.5103 | 0.8644 | 0.5726 | 23.5183 | 4.1317 | 0.0106 | 94.7223 |
| ResNeXt101 Xie et al. (2017) | 0.7463 | 0.5604 | 0.5397 | 0.5326 | 0.8800 | 0.6168 | 42.1389 | 8.0529 | 0.1494 | 6.6929 |
| ShuffleNetV2 Ma et al. (2018) | 0.7632 | 0.5849 | 0.5557 | 0.5580 | 0.8899 | 0.6403 | 0.9217 | 0.1473 | 0.0086 | 116.7053 |
| SimpleViT Beyer et al. (2022) | 0.7196 | 0.5230 | 0.4489 | 0.4391 | 0.8523 | 0.5715 | **0.3808** | 0.0768 | 0.0056 | 177.2153 |
| VAN Guo et al. (2023) | 0.7217 | 0.5207 | 0.4961 | 0.5023 | 0.8501 | 0.5735 | 3.8469 | 0.8706 | 0.0120 | 83.2445 |
| Ours | **0.7848** | **0.6436** | **0.5843** | **0.5977** | **0.8978** | **0.6738** | 2.5623 | 3.3584 | 0.0619 | 16.1646 |
| C: Kvasir Dataset |  |  |  |  |  |  |  |  |  |  |
| Method | Accuracy ↑ | Precision ↑ | Recall ↑ | F1 Score↑ | AUC ↑ | MCC ↑ | Params ↓ | Flops ↓ | Time ↓ | FPS ↓ |
| ConvMixer Trockman and Kolter (2022) | 0.8044 | 0.8089 | 0.8044 | 0.8026 | 0.9717 | 0.7776 | 3.7612 | 3.8609 | 0.0949 | 10.5350 |
| DenseNet121 Huang et al. (2017) | 0.8332 | 0.8403 | 0.8332 | 0.8313 | 0.9821 | 0.8110 | 6.9621 | 2.8960 | 0.0201 | 49.7727 |
| EfficientFormerL1 Li et al. (2022) | 0.7481 | 0.7579 | 0.7481 | 0.7425 | 0.9715 | 0.7149 | 11.3918 | 1.3095 | 0.0475 | 21.0739 |
| EfficientNetB0 Tan and Le (2019) | 0.8029 | 0.8067 | 0.8029 | 0.8012 | 0.9759 | 0.7757 | 4.0178 | 0.4139 | 0.0105 | 94.7937 |
| InceptionNextTiny Yu et al. (2024) | 0.7369 | 0.7397 | 0.7369 | 0.7329 | 0.9617 | 0.7008 | 25.7625 | 4.1982 | 0.0098 | 101.6899 |
| MLPMixer Tolstikhin et al. (2021) | 0.7280 | 0.7299 | 0.7280 | 0.7273 | 0.9543 | 0.6896 | 5.4242 | 1.3170 | **0.0036** | **279.0365** |
| MobileViTXXS Mehta and Rastegari (2021) | 0.7825 | 0.7885 | 0.7825 | 0.7792 | 0.9759 | 0.7531 | 1.0140 | 0.2731 | 0.0118 | 84.8797 |
| MobileNetV3 Howard et al. (2019) | 0.8085 | 0.8139 | 0.8085 | 0.8076 | 0.9786 | 0.7821 | 1.5261 | **0.0615** | 0.0091 | 109.6956 |
| RegionViT Chen et al. (2021) | 0.7310 | 0.7345 | 0.7310 | 0.7281 | 0.9674 | 0.6939 | 12.2376 | 2.2633 | 0.0528 | 18.9278 |
| RepVGG Ding et al. (2021) | 0.8114 | 0.8127 | 0.8114 | 0.8103 | 0.9673 | 0.7849 | 10.2606 | 2.8682 | 0.0431 | 23.2149 |
| Resnet50 He et al. (2016) | 0.7641 | 0.7721 | 0.7641 | 0.7624 | 0.9679 | 0.7320 | 23.5244 | 4.1317 | 0.0109 | 91.4189 |
| ResNeXt101 Xie et al. (2017) | 0.7675 | 0.7797 | 0.7675 | 0.7640 | 0.9620 | 0.7368 | 42.1451 | 8.0529 | 0.1419 | 7.0477 |
| ShuffleNetV2 Ma et al. (2018) | 0.8518 | 0.8540 | 0.8518 | 0.8516 | **0.9854** | 0.8309 | 0.9246 | 0.1473 | 0.0084 | 118.7920 |
| SimpleViT Beyer et al. (2022) | 0.7269 | 0.7329 | 0.7269 | 0.7245 | 0.9651 | 0.6893 | **0.3810** | 0.0768 | 0.0039 | 253.2973 |
| VAN Guo et al. (2023) | 0.7644 | 0.7664 | 0.7644 | 0.7624 | 0.9699 | 0.7315 | 3.8477 | 0.8706 | 0.0113 | 88.2147 |
| Ours | **0.8558** | **0.8650** | **0.8557** | **0.8554** | 0.9850 | **0.8366** | 2.5623 | 3.3584 | 0.0642 | 15.5713 |
| D: Skin-Cancer Dataset |  |  |  |  |  |  |  |  |  |  |
| Method | Accuracy ↑ | Precision ↑ | Recall ↑ | F1 Score↑ | AUC ↑ | MCC ↑ | Params ↓ | Flops ↓ | Time ↓ | FPS ↓ |
| ConvMixer Trockman and Kolter (2022) | 0.5231 | 0.4680 | 0.4623 | 0.4550 | **0.8682** | 0.4425 | 3.7617 | 3.8609 | 0.0849 | 11.7729 |
| DenseNet121 Huang et al. (2017) | 0.5180 | 0.4762 | 0.4489 | 0.4497 | 0.8415 | 0.4363 | 6.9631 | 2.8960 | 0.0199 | 50.2135 |
| EfficientFormerL1 Li et al. (2022) | 0.4888 | 0.3591 | 0.3628 | 0.3314 | 0.8325 | 0.3914 | 11.3927 | 1.3095 | 0.0334 | 29.8995 |
| EfficientNetB0 Tan and Le (2019) | 0.4625 | 0.4008 | 0.3893 | 0.3827 | 0.8427 | 0.3751 | 4.0191 | 0.4139 | 0.0113 | 88.8317 |
| InceptionNextTiny Yu et al. (2024) | 0.4455 | 0.4033 | 0.3813 | 0.3837 | 0.8318 | 0.3501 | 25.7648 | 4.1983 | 0.0091 | 109.8458 |
| MLPMixer Tolstikhin et al. (2021) | 0.4866 | 0.4110 | 0.3895 | 0.3916 | 0.8394 | 0.3954 | 5.4247 | 1.3170 | **0.0039** | **254.3940** |
| MobileViTXXS Mehta and Rastegari (2021) | 0.4968 | 0.4140 | 0.3930 | 0.3720 | 0.8390 | 0.4071 | 1.0144 | 0.2731 | 0.0108 | 92.5576 |
| MobileNetV3 Howard et al. (2019) | 0.4981 | 0.4470 | 0.4346 | 0.4312 | 0.8549 | 0.4137 | 1.5271 | **0.0615** | 0.0090 | 110.6497 |
| RegionViT Chen et al. (2021) | 0.4883 | 0.4035 | 0.4058 | 0.4006 | 0.8337 | 0.3991 | 12.2381 | 2.2633 | 0.0528 | 18.9483 |
| RepVGG Ding et al. (2021) | 0.5180 | 0.4360 | 0.4477 | 0.4326 | 0.8487 | 0.4372 | 10.2611 | 2.8682 | 0.0315 | 31.7892 |
| Resnet50 He et al. (2016) | 0.5096 | 0.4479 | 0.4396 | 0.4367 | 0.8580 | 0.4242 | 23.5265 | 4.1317 | 0.0099 | 100.7933 |
| ResNeXt101 Xie et al. (2017) | 0.4994 | 0.4368 | 0.4123 | 0.4087 | 0.8337 | 0.4132 | 42.1471 | 8.0529 | 0.1411 | 7.0866 |
| ShuffleNetV2 Ma et al. (2018) | 0.5405 | 0.4858 | 0.4764 | 0.4704 | 0.8563 | 0.4635 | 0.9256 | 0.1473 | 0.0070 | 142.5641 |
| SimpleViT Beyer et al. (2022) | 0.4425 | 0.3719 | 0.3678 | 0.3640 | 0.8207 | 0.3456 | **0.3811** | 0.0768 | 0.0055 | 180.6023 |
| VAN Guo et al. (2023) | 0.4981 | 0.4195 | 0.4087 | 0.4060 | 0.8440 | 0.4080 | 3.8479 | 0.8706 | 0.0106 | 94.7108 |
| Ours | **0.6092** | **0.5235** | **0.5304** | **0.5161** | 0.8999 | **0.5408** | 2.5623 | 3.3584 | 0.0677 | 14.7751 |

#### 4.3.2. Qualitative Performance

In addition to quantitative evaluation, we conduct qualitative experiments to assess the interpretability and uncertainty estimation of our proposed method. While quantitative metrics demonstrate classification accuracy, qualitative results provide deeper insights into the model’s decision-making process and reliability in clinical applications.

##### Uncertainty Maps

To demonstrate uncertainty quantification, we generate pixel-level uncertainty maps using our model. These maps reveal regions where the model is less confident. For example, in diabetic retinopathy images, uncertainty tends to increase in poorly illuminated or blurred regions. Such visual cues provide clinicians with valuable information regarding the trustworthiness of predictions.

In order to better understand the reliability of the proposed model, we further analyze its predictive uncertainty on the Skin-Cancer dataset. Fig. 3 illustrates the five most uncertain and the five most certain cases as estimated by our framework. The most certain cases demonstrate high-confidence predictions that align with the ground-truth labels, highlighting the model’s robustness in handling clear diagnostic patterns. In contrast, the most uncertain cases correspond to challenging samples, such as those with ambiguous lesion boundaries or visually similar inter-class characteristics, where the model exhibits lower confidence. This analysis not only emphasizes the importance of incorporating uncertainty estimation in medical image classification but also provides a valuable diagnostic aid for identifying cases that may require additional expert review.

**Figure 3.**
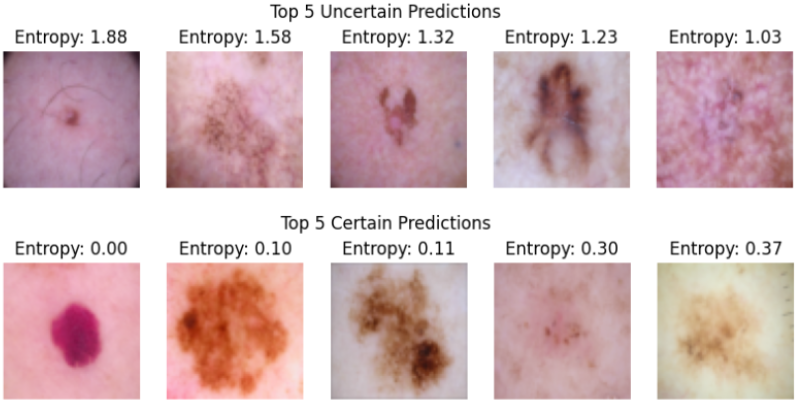
Illustration of the 5 most uncertain and the 5 most certain cases using the skin cancer dataset.

To further investigate the interpretability of our framework, we visualize the GradCAM-based attention maps along with uncertainty quantification on the Skin Cancer dataset, as shown in Fig. 4.

**Figure 4.**
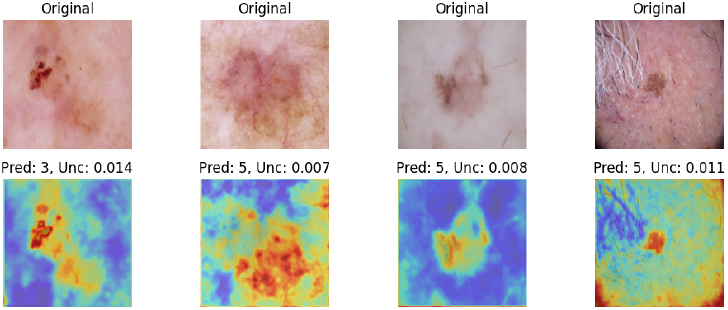
Illustration of GradCAM-based attention maps along with predicted labels and uncertainty quantification using the skin cancer dataset.

Figures 5, 6, and 7 provide qualitative insights into the model’s performance. Figure 5 shows the confusion matrix for one fold, illustrating that the classifier achieves high true positive rates across most classes with minimal misclassifications. Figure 6 presents the ROC-AUC curve for another fold, demonstrating strong discriminative capability between classes, with most curves approaching the ideal top-left corner. Figure 7 displays the precision-recall curves, highlighting that the model maintains high precision even at elevated recall levels, indicating reliable performance on imbalanced class distributions.

**Figure 5.**
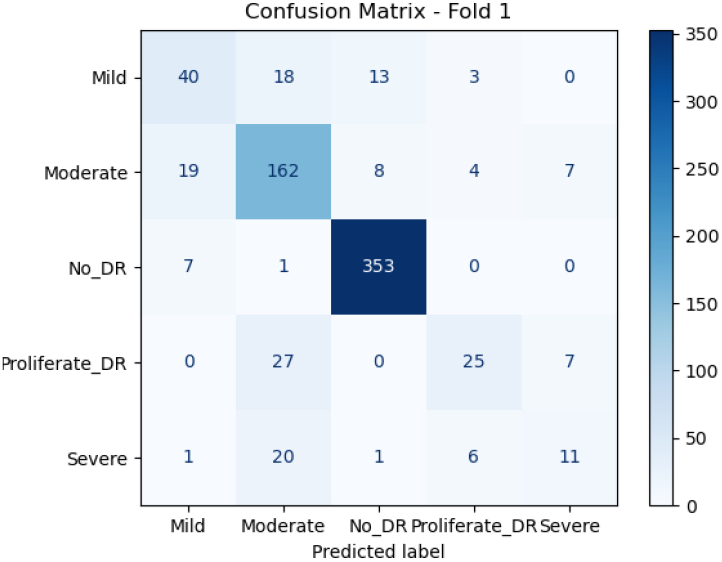
Confusion matrix using the diabetic retinopathy dataset.

**Figure 6.**
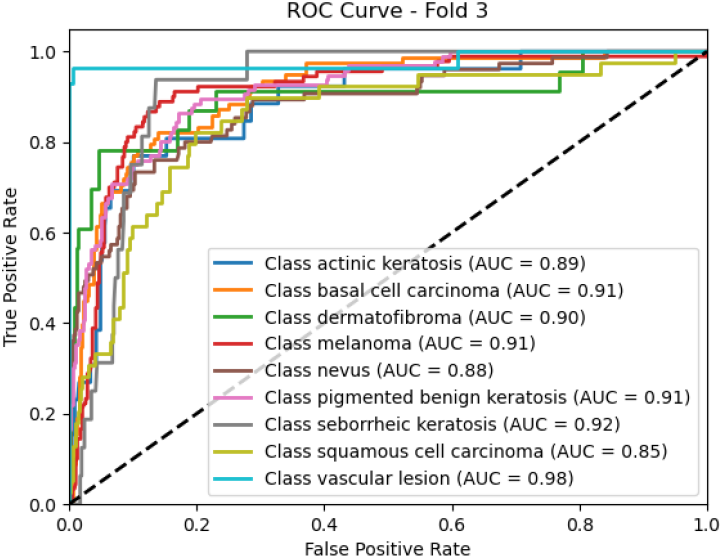
ROC-AUC curve using the skin cancer dataset.

**Figure 7.**
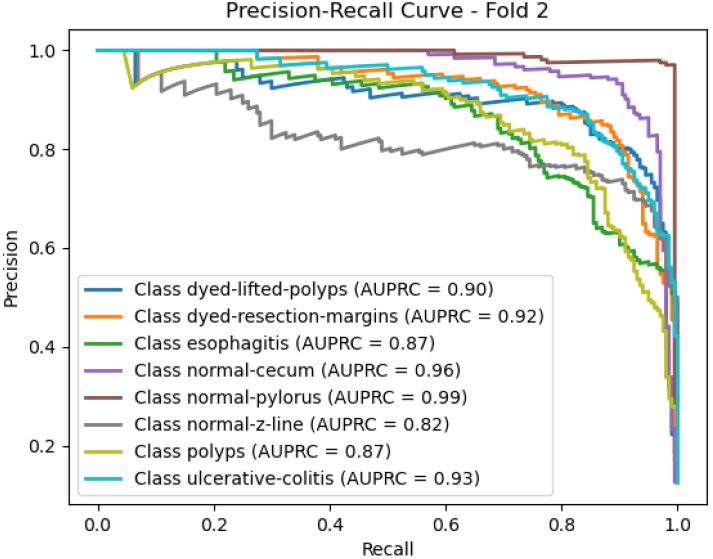
Precision-Recall curve using the Kvasir dataset.

##### Attention Visualization

We visualize the attention maps generated using the GradCAM technique. These visualizations highlight the most discriminative regions in the input medical images that contribute to the classification decision. For instance, in skin lesion images, the attention maps emphasize irregular pigmentation areas that are clinically relevant. This confirms that our attention-enhanced Bayesian model aligns well with human expert reasoning.

###### Reliability Diagram

To evaluate the calibration performance of our model, we present the reliability diagram on the Diabetic Retinopathy dataset in Fig. 9. The diagram compares the predicted confidence levels against the observed accuracy, offering insights into how well the model’s probabilistic outputs align with actual outcomes. An ideally calibrated model would follow the diagonal line, where confidence perfectly matches accuracy. Our method demonstrates a closer alignment to this diagonal compared to conventional baselines, indicating reduced overconfidence and improved trustworthiness of predictions. Such well-calibrated probabilistic estimates are particularly important in clinical practice, where decision-making depends not only on classification accuracy but also on the reliability of the confidence scores provided by the model.

**Figure 8.**
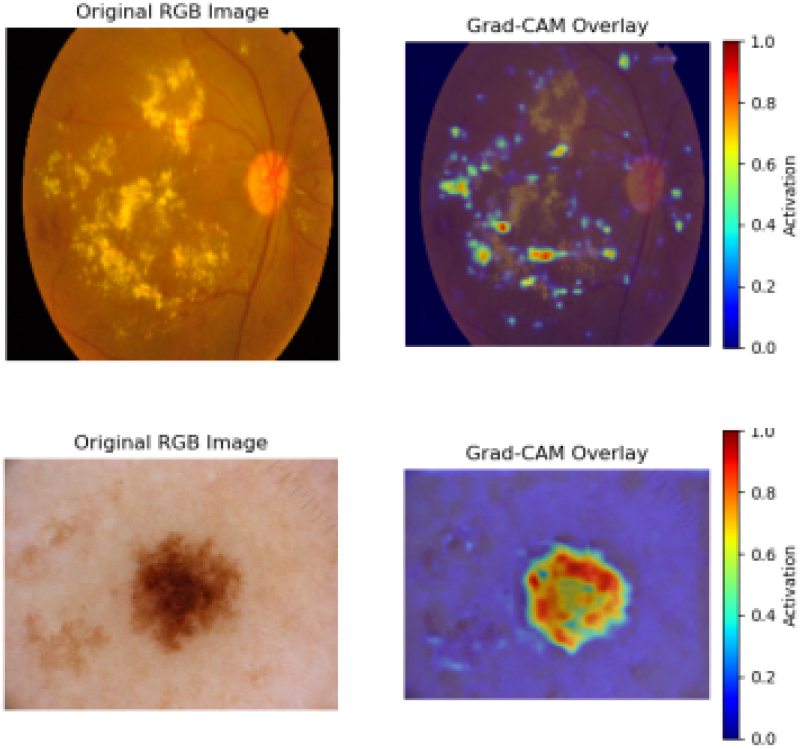
GradCAM visualization using the diabetic retinopathy dataset on the top row and skin cancer dataset on the bottom row.

**Figure 9.**
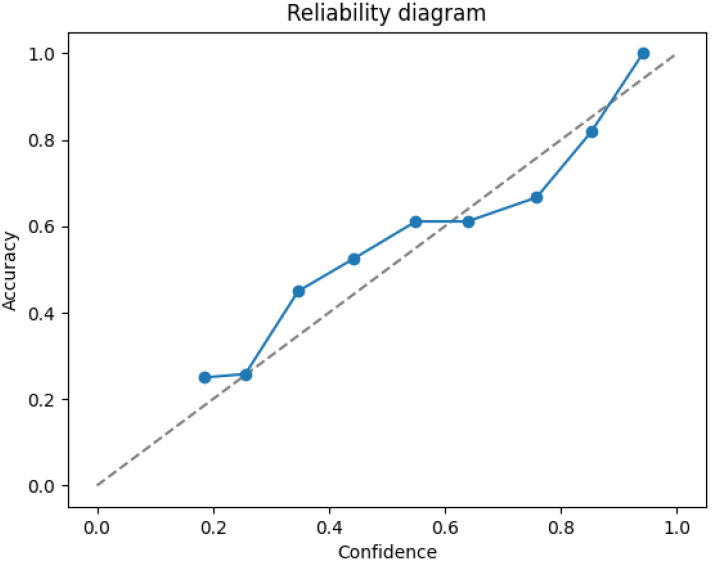
Reliability Diagram using the Diabetic Retinopathy dataset. The Y-axis denotes accuracy, and the X-axis denotes confidence.

### 4.4. Ablation Study

The ablation studies summarized in Table 2 and Table 3 respectively, provide a detailed assessment of the contribution of individual components and loss functions in our framework. In the architectural ablations, removing the spiral module led to a noticeable drop in recall and F1, while eliminating the squeeze block reduced both calibration metrics (Brier and ECE), highlighting their complementary role in performance. Replacing Bayesian components with fully connected (BNN → FC) or Monte Carlo (BNN → MC) variants showed moderate gains in precision or recall but sacrificed overall stability in calibration. In contrast, our full model consistently achieved the best balance across accuracy (0.6092), recall (0.5304), F1 score (0.5161), MCC (0.5408), and calibration (Brier 0.1239, ECE 0.0568), demonstrating the synergistic effect of all components. Similarly, the loss function analysis confirms that our proposed approach provides consistent improvements over the standard cross-entropy baseline. While CE achieved reasonable performance, our method delivered higher values across all metrics, including a 2.8% gain in accuracy and 4.1% in MCC, while also lowering calibration errors. These results collectively validate the design choices of our framework and emphasize the importance of both architectural innovations and uncertainty-aware loss design for reliable medical image classification.

**Table 2:** Ablation study using different components in the network architecture. The best values are highlighted in bold.

| Method | Acc ↑ | Pre ↑ | Rec ↑ | F1 ↑ | AUC ↑ | MCC ↑ | Brier ↓ | ECE ↓ | Param ↓ | Flops ↓ | Time ↓ | FPS ↓ |
| --- | --- | --- | --- | --- | --- | --- | --- | --- | --- | --- | --- | --- |
| W/o Spiral | 0.5914 | 0.5297 | 0.5099 | 0.5083 | 0.8877 | 0.5209 | 0.1351 | 0.0598 | **2.4101** | **3.1529** | **0.0101** | **99.0292** |
| W/o Squeeze | 0.6058 | 0.5208 | 0.4992 | 0.4930 | **0.9087** | 0.5347 | 0.1264 | 0.0665 | 2.5427 | 3.3568 | 0.0693 | 14.4231 |
| BNN→FC | 0.5995 | **0.5370** | 0.5011 | 0.4998 | 0.9045 | 0.5287 | 0.1270 | 0.0625 | 2.5636 | 3.3584 | 0.0707 | 14.1350 |
| BNN→MC | 0.6046 | 0.5310 | 0.5200 | 0.5120 | 0.8978 | 0.5368 | 0.1410 | 0.0587 | 2.5636 | 3.3584 | 0.0701 | 14.2669 |
| Ours | **0.6092** | 0.5235 | **0.5304** | **0.5161** | 0.8999 | **0.5408** | **0.1239** | **0.0568** | 2.5623 | 3.3584 | 0.0677 | 14.7751 |

**Table 3:** Ablation study using loss function. The best values are highlighted in bold.

| Method | Acc ↑ | Pre ↑ | Rec ↑ | F1 ↑ | AUC ↑ | MCC ↑ | Brier ↓ | ECE ↓ |
| --- | --- | --- | --- | --- | --- | --- | --- | --- |
| CE | 0.5810 | 0.5023 | 0.5150 | 0.5081 | 0.8912 | 0.5001 | 0.1256 | 0.0623 |
| Ours | **0.6092** | **0.5235** | **0.5304** | **0.5161** | **0.8999** | **0.5408** | **0.1239** | **0.0568** |

### 4.5. Failure Cases

Despite the overall strong performance of our proposed Bayesian Neural Network with channel and spiral attention, certain failure cases were observed during experiments. Understanding these failure modes is important for further improvement and clinical applicability.

1. **Low-Quality or Noisy Images:** Medical images with low resolution, poor contrast, or significant noise often lead to misclassifications. For example, blurry retinal fundus images in the Diabetic Retinopathy dataset or poorly illuminated skin lesion images sometimes result in incorrect predictions. While the model indicates higher uncertainty in these cases, it still occasionally misclassifies the images, highlighting limitations in handling severely degraded inputs.
2. **Rare or Ambiguous Classes:** Classes with very few training samples, such as certain less frequent skin lesion categories, are more prone to misclassification. The model tends to be overconfident in some of these rare categories, even though Bayesian modeling partially mitigates this effect. This underscores the importance of balanced datasets for reliable uncertainty estimation.
3. **Subtle or Small Lesions:** In cases where pathological regions are extremely small or visually subtle, spiral attention may fail to fully capture the critical areas. For instance, small microaneurysms in retinal images or tiny early-stage skin lesions can be overlooked, leading to incorrect predictions. These failure cases often correlate with higher predictive entropy, signaling that uncertainty estimates are still informative.
4. **Confounding Visual Features:** Occasionally, non-pathological features, such as artifacts, markings, or anatomical variations, can be incorrectly attended to by the network. This can mislead both the attention modules and the classifier, resulting in errors. In such cases, uncertainty estimates tend to be moderate but not always sufficient to flag potential misclassifications.

## 5. Conclusions

In this work, we proposed a novel Bayesian deep learning framework for multi-class image classification. The architecture integrates residual blocks with Squeeze-and-Excitation (SE) modules and Spiral Attention, combined with a Bayes-by-Backprop linear head to capture epistemic uncertainty. Quantitative results demonstrate that our approach achieves high classification accuracy while maintaining well-calibrated probability predictions as indicated by a low ECE. Furthermore, the incorporation of Bayesian modeling allows the model to estimate epistemic uncertainty, providing interpretable confidence measures alongside predictions. This capability is particularly beneficial in clinical and safety-critical applications, where understanding the certainty of predictions is crucial. Future work may explore extending the architecture to larger datasets, multi-modal inputs, and real-time deployment scenarios.

## Data Availability

All data produced are available online at

https://www.kaggle.com/datasets/sartajbhuvaji/brain-tumor-classification-mri

## References

Aalishah, R., Navardi, M., Mohsenin, T., 2025. Medmambalite: Hardware-aware mamba for medical image classification. arXiv preprint 2508.05049 .

Abdar, M., Fahami, M.A., Chakrabarti, S., Khosravi, A., Pławiak, P., Acharya, U.R., Tadeusiewicz, R., Nahavandi, S., 2021. Barf: A new direct and cross-based binary residual feature fusion with uncertainty-aware module for medical image classification. Information Sciences 577, 353–378.

Abdar, M., Fahami, M.A., Rundo, L., Radeva, P., Frangi, A.F., Acharya, U.R., Khosravi, A., Lam, H.K., Jung, A., Nahavandi, S., 2022. Hercules: Deep hierarchical attentive multilevel fusion model with uncertainty quantification for medical image classification. IEEE Transactions on Industrial Informatics 19, 274–285.

Almalik, F., Yaqub, M., Nandakumar, K., 2022. Self-ensembling vision transformer (sevit) for robust medical image classification, in: International conference on medical image computing and computer-assisted intervention, Springer. pp. 376–386.

Ansari, S.A., Agrawal, A.P., Wajid, M.A., Wajid, M.S., Zafar, A., 2024. Metav: A pioneer in feature augmented meta-learning based vision transformer for medical image classification. Interdisciplinary Sciences: Computational Life Sciences 16, 469–488.

Asia Pacific Tele-Ophthalmology Society (APTOS), 2019. Aptos 2019 blindness detection. https://www.kaggle.com/competitions/aptos2019-blindness-detection. Kaggle dataset.

Beyer, L., Zhai, X., Kolesnikov, A., 2022. Better plain vit baselines for imagenet-1k. arXiv preprint 2205.01580 .

Cai, Z., Chen, Y., Wang, J., He, X., Pei, Z., Lei, X., Lu, C., 2025. Dafnet: A novel dynamic adaptive fusion network for medical image classification. Information Fusion, 103507.

Chen, C.F., Panda, R., Fan, Q., 2021. Regionvit: Regional-to-local attention for vision transformers. arXiv preprint 2106.02689.

Cheng, J., Tian, S., Yu, L., Gao, C., Kang, X., Ma, X., Wu, W., Liu, S., Lu, H., 2022. Resganet: Residual group attention network for medical image classification and segmentation. Medical Image Analysis 76, 102313.

Chowdary, G.J., Yin, Z., 2024. Med-former: A transformer based architecture for medical image classification, in: International conference on medical image computing and computer-assisted intervention, Springer. pp. 448–457.

Codella, N., Rotemberg, V., Tschandl, P., Celebi, M.E., Dusza, S., Gutman, D., Helba, B., Kalloo, A., Liopyris, K., Marchetti, M., et al., 2019. Skin lesion analysis toward melanoma detection 2018: A challenge hosted by the international skin imaging collaboration (isic). arXiv preprint 1902.03368 .

Dai, Y., Gao, Y., Liu, F., 2021. Transmed: Transformers advance multi-modal medical image classification. Diagnostics 11, 1384.

Ding, X., Zhang, X., Ma, N., Han, J., Ding, G., Sun, J., 2021. Repvgg: Making vgg-style convnets great again, in: Proceedings of the IEEE/CVF conference on computer vision and pattern recognition, pp. 13733–13742.

Djoumessi, K., Mensah, S.O., Berens, P., 2025. A hybrid fully convolutional cnn-transformer model for inherently interpretable medical image classification. arXiv preprint 2504.08481 .

Dosovitskiy, A., Beyer, L., Kolesnikov, A., Weissenborn, D., Zhai, X., Unterthiner, T., Dehghani, M., Minderer, M., Heigold, G., Gelly, S., et al., 2020. An image is worth 16x16 words: Transformers for image recognition at scale. arXiv preprint 2010.11929 .

Fang, L., Wang, C., Li, S., Rabbani, H., Chen, X., Liu, Z., 2019. Attention to lesion: Lesion-aware convolutional neural network for retinal optical coherence tomography image classification. IEEE transactions on medical imaging 38, 1959–1970.

Gal, Y., Ghahramani, Z., 2016. Dropout as a bayesian approximation: Representing model uncertainty in deep learning, in: international conference on machine learning, PMLR. pp. 1050–1059.

Galdran, A., Carneiro, G., González Ballester, M.A., 2021. Balanced-mixup for highly imbalanced medical image classification, in: International Conference on Medical Image Computing and Computer-Assisted Intervention, Springer. pp. 323–333.

Gao, L., Zhang, L., Liu, C., Wu, S., 2020. Handling imbalanced medical image data: A deep-learning-based one-class classification approach. Artificial intelligence in medicine 108, 101935.

Guo, M.H., Lu, C.Z., Liu, Z.N., Cheng, M.M., Hu, S.M., 2023. Visual attention network. Computational visual media 9, 733–752.

He, K., Zhang, X., Ren, S., Sun, J., 2016. Deep residual learning for image recognition, in: Proceedings of the IEEE conference on computer vision and pattern recognition, pp. 770–778.

He, X., Deng, Y., Fang, L., Peng, Q., 2021. Multi-modal retinal image classification with modality-specific attention network. IEEE transactions on medical imaging 40, 1591–1602.

Howard, A., Sandler, M., Chu, G., Chen, L.C., Chen, B., Tan, M., Wang, W., Zhu, Y., Pang, R., Vasudevan, V., et al., 2019. Searching for mobilenetv3, in: Proceedings of the IEEE/CVF international conference on computer vision, pp. 1314–1324.

Hu, J., Shen, L., Sun, G., 2018. Squeeze-and-excitation networks, in: Proceedings of the IEEE conference on computer vision and pattern recognition, pp. 7132–7141.

Hu, J., Xiang, Y., Lin, Y., Du, J., Zhang, H., Liu, H., 2025. Multi-scale transformer architecture for accurate medical image classification, in: Proceedings of the 2025 International Conference on Artificial Intelligence and Computational Intelligence, pp. 409–414.

Huang, G., Liu, Z., Van Der Maaten, L., Weinberger, K.Q., 2017. Densely connected convolutional networks, in: Proceedings of the IEEE conference on computer vision and pattern recognition, pp. 4700–4708.

Huix, J.P., Ganeshan, A.R., Haslum, J.F., Söderberg, M., Matsoukas, C., Smith, K., 2024. Are natural domain foundation models useful for medical image classification?, in: Proceedings of the IEEE/CVF winter conference on applications of computer vision, pp. 7634– 7643.

Hussain, T., Shouno, H., Hussain, A., Hussain, D., Ismail, M., Mir, T.H., Hsu, F.R., Alam, T., Akhy, S.A., 2025. Effresnet-vit: A fusion-based convolutional and vision transformer model for explainable medical image classification. IEEE Access .

Hussain, Z., Gimenez, F., Yi, D., Rubin, D., 2018. Differential data augmentation techniques for medical imaging classification tasks, in: AMIA annual symposium proceedings, p. 979.

Ju, L., Wang, X., Wang, L., Mahapatra, D., Zhao, X., Zhou, Q., Liu, T., Ge, Z., 2022. Improving medical images classification with label noise using dual-uncertainty estimation. IEEE transactions on medical imaging 41, 1533–1546.

Li, J., Feng, M., Xia, C., 2024a. Dbcvt: Double branch convolutional transformer for medical image classification. Pattern Recognition Letters 186, 250–257.

Li, Y., Huang, Y., He, N., Ma, K., Zheng, Y., 2024b. Improving vision transformer for medical image classification via token-wise perturbation. Journal of Visual Communication and Image Representation 98, 104022.

Li, Y., Yuan, G., Wen, Y., Hu, J., Evangelidis, G., Tulyakov, S., Wang, Y., Ren, J., 2022. Efficientformer: Vision transformers at mobilenet speed. Advances in Neural Information Processing Systems 35, 12934–12949.

Liang, G., Zhang, Y., Wang, X., Jacobs, N., 2020. Improved trainable calibration method for neural networks on medical imaging classification. arXiv preprint 2009.04057 .

Liu, S., Wang, L., Yue, W., 2024. An efficient medical image classification network based on multi-branch cnn, token grouping transformer and mixer mlp. Applied Soft Computing 153, 111323.

Ma, N., Zhang, X., Zheng, H.T., Sun, J., 2018. Shufflenet v2: Practical guidelines for efficient cnn architecture design, in: Proceedings of the European conference on computer vision (ECCV), pp. 116–131.

Manzari, O.N., Ahmadabadi, H., Kashiani, H., Shokouhi, S.B., Ayatollahi, A., 2023. Medvit: a robust vision transformer for generalized medical image classification. Computers in biology and medicine 157, 106791.

Mehta, S., Rastegari, M., 2021. Mobilevit: light-weight, general-purpose, and mobile-friendly vision transformer. arXiv preprint 2110.02178 .

Okolo, G.I., Katsigiannis, S., Ramzan, N., 2022. Ievit: An enhanced vision transformer architecture for chest x-ray image classification. Computer Methods and Programs in Biomedicine 226, 107141.

Pogorelov, K., Randel, K.R., Griwodz, C., Eskeland, S.L., de Lange, T., Johansen, D., Spampinato, C., Dang-Nguyen, D.T., Lux, M., Schmidt, P.T., et al., 2017. Kvasir: A multi-class image dataset for computer aided gastrointestinal disease detection, in: Proceedings of the 8th ACM on Multimedia Systems Conference, pp. 164–169.

Raghu, M., Zhang, C., Kleinberg, J., Bengio, S., 2019. Transfusion: Understanding transfer learning for medical imaging. Advances in neural information processing systems 32.

Rajaraman, S., Ganesan, P., Antani, S., 2022. Deep learning model calibration for improving performance in class-imbalanced medical image classification tasks. PloS one 17, e0262838.

Sagar, A., 2025. Bucan: Bayesian uncertainty-aware classification with attention networks for medical images. medRxiv, 2025–11.

Shaik, N.S., Cherukuri, T.K., Veeranjaneulu, N., Bodapati, J.D., 2024. Medtransnet: advanced gating transformer network for medical image classification. Machine Vision and Applications 35, 73.

Shastri, S., Kansal, I., Kumar, S., Singh, K., Popli, R., Mansotra, V., 2022. Cheximagenet: a novel architecture for accurate classification of covid-19 with chest x-ray digital images using deep convolutional neural networks. Health and technology 12, 193–204.

Sherkatghanad, Z., Abdar, M., Bakhtyari, M., Pławiak, P., Makarenkov, V., 2025. Baytta: Uncertainty-aware medical image classification with optimized test-time augmentation using bayesian model averaging. Knowledge-Based Systems, 114123.

Swati, Z.N.K., Zhao, Q., Kabir, M., Ali, F., Ali, Z., Ahmed, S., Lu, J., 2019. Brain tumor classification for mr images using transfer learning and fine-tuning. Computerized Medical Imaging and Graphics 75, 34–46.

Tajbakhsh, N., Shin, J.Y., Gurudu, S.R., Hurst, R.T., Kendall, C.B., Gotway, M.B., Liang, J., 2016. Convolutional neural networks for medical image analysis: Full training or fine tuning? IEEE transactions on medical imaging 35, 1299–1312.

Tan, M., Le, Q., 2019. Efficientnet: Rethinking model scaling for convolutional neural networks, in: International conference on machine learning, PMLR. pp. 6105–6114.

Tolstikhin, I.O., Houlsby, N., Kolesnikov, A., Beyer, L., Zhai, X., Unterthiner, T., Yung, J., Steiner, A., Keysers, D., Uszkoreit, J., et al., 2021. Mlp-mixer: An all-mlp architecture for vision. Advances in neural information processing systems 34, 24261–24272.

Trockman, A., Kolter, J.Z., 2022. Patches are all you need? arXiv preprint 2201.09792 .

Wang, X., Yang, S., Zhang, J., Wang, M., Zhang, J., Yang, W., Huang, J., Han, X., 2022. Transformer-based unsupervised contrastive learning for histopathological image classification. Medical image analysis 81, 102559.

Wen, Y., Chen, B., Shi, W., Feng, D., Cao, W., Wu, S., 2025. Mspfm: Multi-scale pyramid fusion mamba for medical image classification: Y. wen et al. The Visual Computer, 1–16.

Wu, X., Feng, Y., Xu, H., Lin, Z., Chen, T., Li, S., Qiu, S., Liu, Q., Ma, Y., Zhang, S., 2023. Ctranscnn: Combining transformer and cnn in multilabel medical image classification. Knowledge-Based Systems 281, 111030.

Wu, X., Gou, G., 2025. Uncertainty bidirectional guidance of multitask mamba network for medical image classification and segmentation. Signal, Image and Video Processing 19, 29.

Xie, S., Girshick, R., Dollár, P., Tu, Z., He, K., 2017. Aggregated residual transformations for deep neural networks, in: Proceedings of the IEEE conference on computer vision and pattern recognition, pp. 1492–1500.

Xie, Y., Richmond, D., 2018. Pre-training on grayscale imagenet improves medical image classification, in: Proceedings of the European conference on computer vision (ECCV) workshops, pp. 0–0.

Xue, C., Dou, Q., Shi, X., Chen, H., Heng, P.A., 2019. Robust learning at noisy labeled medical images: Applied to skin lesion classification, in: 2019 IEEE 16th International symposium on biomedical imaging (ISBI 2019), IEEE. pp. 1280–1283.

Xue, C., Yu, L., Chen, P., Dou, Q., Heng, P.A., 2022. Robust medical image classification from noisy labeled data with global and local representation guided co-training. IEEE transactions on medical imaging 41, 1371–1382.

Yang, Y., Fu, H., Aviles-Rivero, A.I., Schönlieb, C.B., Zhu, L., 2023. Diffmic: Dual-guidance diffusion network for medical image classification, in: International conference on medical image computing and computer-assisted intervention, Springer. pp. 95–105.

Yu, W., Zhou, P., Yan, S., Wang, X., 2024. Inceptionnext: When inception meets convnext, in: Proceedings of the IEEE/cvf conference on computer vision and pattern recognition, pp. 5672–5683.

Yue, Y., Li, Z., 2024. Medmamba: Vision mamba for medical image classification. arXiv preprint 2403.03849 .

Zhou, L., Liu, H., Bae, J., He, J., Samaras, D., Prasanna, P., 2023. Self pre-training with masked autoencoders for medical image classification and segmentation, in: 2023 IEEE 20th international symposium on biomedical imaging (ISBI), IEEE. pp. 1–6.

